# Tumor-infiltrating immune cells signature predicts recurrence free survival after complete resection of localized primary gastrointestinal stromal tumors

**DOI:** 10.1101/2020.02.28.20025908

**Authors:** Mengshi Yi, Rui Zhao, Qianyi Wan, Xiaoting Wu, Wen Zhuang, Hanshuo Yang, Chuncheng Wu, Lin Xia, Yi Chen, Yong Zhou

## Abstract

**Background:** Growing evidence has proposed prognostic value of immune infiltration in gastrointestinal stromal tumors (GISTs). Therefore, we aimed to develop a novel immune-based prognostic classifier (IPC) to help better stratify and predict prognosis of GISTs.

**Methods:** The gene expression profiles of 22 immune features of GISTs were detected from GEO dataset. The IPC was constructed using the LASSO Cox regression model and validated in a cohort including 54 patients with complete resection of localized primary GISTs via immunohistochemistry process. The performance assessment of the IPC was estimated, then compared with conventional risk prognostic criteria.

**Results:** The IPC was established based on 4 features: CD8, CD8/CD3, CD68, CD163/CD68 and validated to be an independent predictor of RFS for GISTs (HR 5.2, 95%CI 1.99-13.65). Significant differences were found between low- and high-IPC group in 5-year RFS (92.6% vs 48.1%, *p* < 0.001). Using the IPC, the high-risk group of the Modified NIH classification was split into two groups in 5-year RFS (low-IPC vs high-IPC, 85.7% vs 30.0%, *p* < 0.001). The IPC showed a higher net benefit than both “treat all” or “treat none” methods for the threshold probability within a range of 0-0.62 and exhibited a performance (AUC 0.842) superior to modified NIH classification (AUC 0.763).

**Conclusion:** The IPC was effective to predict RFS after complete resection of localized primary GISTs, adding prognostic value to the routine clinical prognostic criteria. Prospective studies are needed to further validate the analytical accuracy and practicability of the IPC in estimating prognosis of GISTs.

## Introduction

Gastrointestinal stromal tumors (GISTs) are the most mesenchymal tumors of the gastrointestinal tract,^1^ with varying malignancy potential ranging from virtually indolent tumors to rapid progressing tumors.^2^ The malignant potential of GIST is not completely derived from the genetic instability from the carcinogenic gene mutation.^3^ It has been established that the tumor progression is a Darwinian evolutionary process, which involved the interplay between cancer subclones and the local immunological microenvironment.^4^ Not only the genetic instability, but also the immune microenvironment play an indispensable role in the malignant progress of GIST.^5^

Recent evidence has suggested the presence of immune infiltration in GISTs,^6^ of which the tumor-associated macrophages (TAMs) were observed the most numerous tumor-infiltrating immune cells,^7,8^ the tumor-infiltrating lymphocytes (TILs) the next.^6,9^ Previous studies have identified the prognostic role of various immune components including CD3+ TILs,^6^ natural killer (NK) cells^6^ and so on. Our previous study proved that the rescue of exhausted CD8+ T cells prolonged survival through blockade of PD-1/PD-L1.^10^ *Maud Toulmond et al* found that the immunosuppression of tumor microenvironment, as a result from macrophage infiltration,^11^ may be related to the activity of PD-1 inhibition in part of GISTs. Nevertheless, it has been not assessed about the predicting potential of comprehensive infiltrating immune cells for GISTs. It is well known that the immune response is involved by different types of cell, myeloid cells, cytotoxic lymphocytes, antigen-presenting cells, for example. Enumerating the immune components based on their specialized markers using computer-based analysis might be useful for promoting studies about the complicated immune infiltration in GISTs and management of its future application in clinical works. The immune signatures observed in association with the boarder phenomenon of immune-mediated, tissue-specific destruction could be summarized as the immune contexture.^12^ In recent years, on the basis of the immune contexture, a comprehensive, standardized, powerful immunoscore system was proposed to make more accurate prognoses to various types of cancers. An international multi-nation corporation of 14 centers assessed the the immunoscore based on the amounts of CD3+ and CD8+ T cells in TNM stage I-III colon cancer, and finally proved that the immunoscore provided the highest relative contribution to the risk of recurrence in colon cancer.^13^ The proposal and effectiveness of the immunoscore has been validated in several other cancers such as gastric cancer,^14^ hepatocellular carcinoma^15^, urothelial carcinoma of the bladder^16^ and so on. These existing evidence indicated that immunoscore provides a more accurate prognosis.^17^

Therefore, we firstly established a novel immune-based prognostic classifier (IPC) for GISTs using the least absolute shrinkage and selection operator method (LASSO) Cox regression model, then evaluated the prognostic performance of the classifier on surgical specimens of patients with clinically localized primary GISTs treated by complete surgery, finally compared the prognostic accuracy of the IPC with current widely used the modified National Institutes of Health (NIH) classification system^18^ and heat maps by *Joensuu*^19^.

## Materials and Methods

### Genomic data from GEO dataset

This study partially made use of data in the public domain. Gene expression data of immune infiltrating cells for patients diagnosed with GISTs were downloaded from GEO dataset (GES8167/GSE47911). All patients with gene expression data and data of clinical outcome from GISTs were considered eligible, with no specific exclusion applied. We waived additional informed consent because these data used in this study were obtained from public databases. Participants in the original genomic studies provided informed consent.

### Development of the immunoscore system

CIBERSORT is a recently introduced computational algorithm for enumeration of different subsets of immune cells using RNA specimens of numerous tissue types (https://cibersort.stanford.edu).^20^ The estimation of the relative fractions of 22 immune cells from expression profiles of GISTs tissues was conducted by CIBERSORT. To construct a new multi-immune feature classifier to predict prognosis of GISTs (IPC), hierarchical clustering was performed by X-cell using GEO dataset. The LASSO Cox analysis, a widely applicable method for regression with high-dimensional prognostic factors was conducted to identify the most useful prognostic immune features and construct this Immunoscore system for GIST on the basis of recurrence free survival (RFS) using the “glment” package.

### Validation dataset

Data collection was conducted from patients with localized primary GISTs who underwent complete resection between July 2012 and June 2014 at Department of Gastrointestinal Surgery, West China Hospital. Patients were included in this research according to the following criteria: complete resection of tumor, pathological confirmed as GISTs, availability of clinicopathological characteristics, follow-up data and tumor samples, without preoperative or postoperative adjuvant imatinib therapy, and absence of any other malignancies or any evidence of metastasis/recurrence at the diagnosis. The institutional review board approved the retrospective analysis of anonymous data.

### Immunohistochemistry and evaluation of immunohistochemical staining

Immunohistochemistry (IHC) process of tumor tissues from the validation cohort was assigned to evaluate the performance of the IPC. To evaluate the infiltrating of immune features in GISTs, the tissue sections were firstly screened at low power (×200) using an inverted research microscope (model DM IRB; Leica, Germany). Five most independent and representative areas were chosen using Leica Qwin Plus v3 software to ensure its representativeness and homogeneity and then photographed at ×400 magnification. Settings used in the process of each photograph were identical. High-resolution spot images (1360 × 1024) were obtained, stored, and then analyzed using a computer-automated method (Image-pro plus 6.0, Media Cybernetics Inc.).^21^ Two independent pathologists blinded to the clinical characteristics and prognosis of GISTs estimated the IHC results of all samples, got the results in agreement in approximately 90% of the cases and if inconsistent, consulted the third pathologist. The value of each feature was recorded, and mean value of the 5 representative fields were used for statistical analysis.

### Statistical analysis

RFS was calculated from surgery till any recurrence, metastasis, death or the deadline date of last follow-up visit (June 2019). Survival analysis was performed using the Kaplan-Meier method and assessed by a two tailed log-rank test. Univariate and multivariate Cox proportional hazard model was used to identify independent prognostic factors, and hazard ratio (HR) was estimated with 95% confidence interval (CI) limits. The prognostic accuracy of the IPC was estimated and compared with the modified NIH risk classification and heat maps by *Joensuu* by conducting receiver operating characteristic (ROC) analysis. Decision curve analysis (DCA) using the “rmda” package was performed to evaluate the clinical utility of the IPC. All statistical analyses were performed using R software (version 3.1.0) and SPSS software (version 12.0). P value were set at 0.05 with statistical significance.

## Results

The workflow was presented in **Figure 1**.

**Figure 1.**
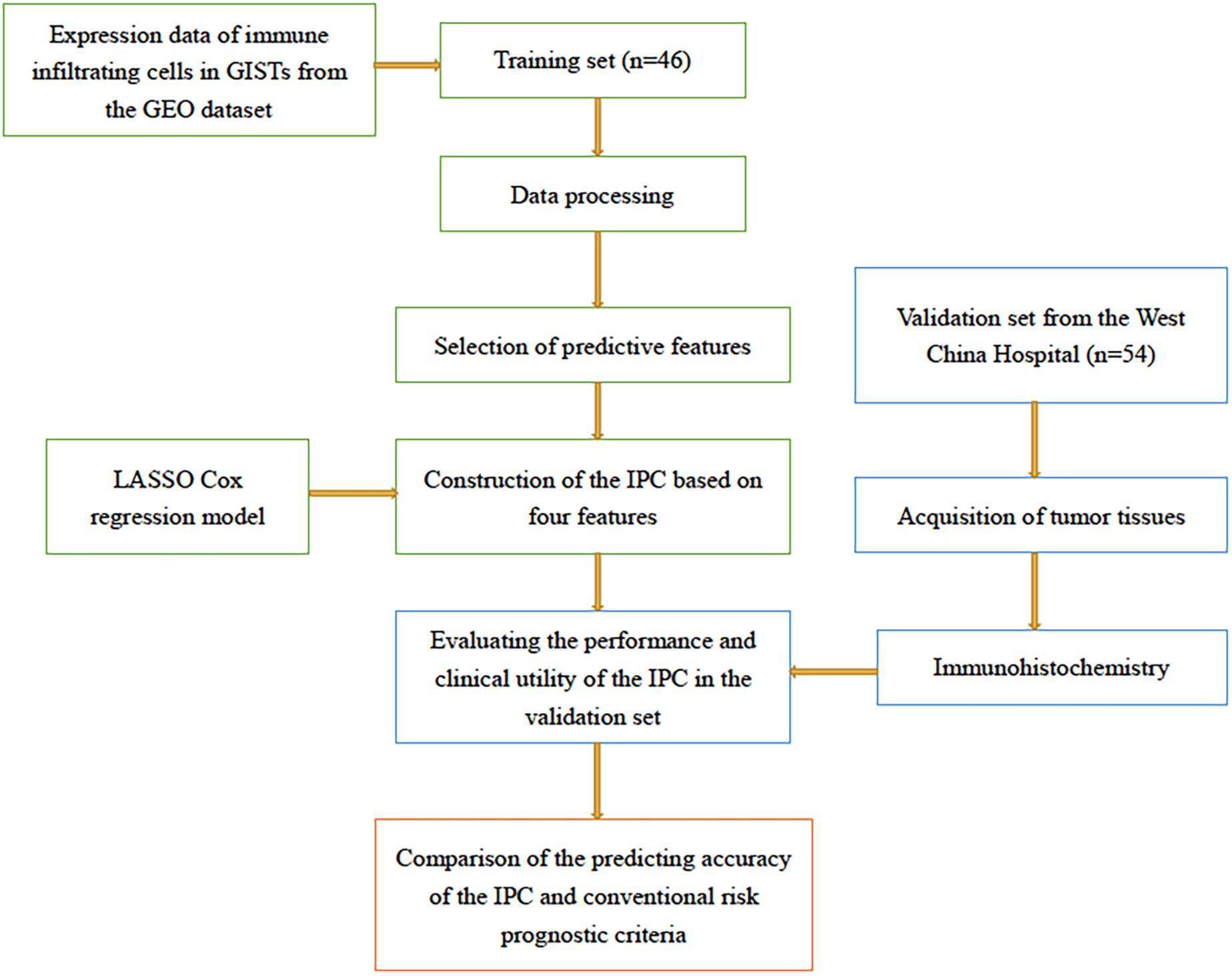
The flowchart of this study.

### Construction of the risk score

Relative fractions of 22 immune cell categories in samples from the training cohort in clusters with non-recurrence and clusters with recurrence was showed in **Figure 2a**. Comparison of relative fractions of these 22 immune cell categories in the two clusters showed a distinct difference in B cells, CD4+ T cells, CD8+ T cells, eosinophils, macrophages, memory B cells, monocytes, neutrophils, T Gamma/Delta cells (Tgd cells), T helper 2 cells (Th2 cells), dendritic cells (DCs) (**Figure 2b and c**). Hierarchical clustering analysis that classified the patients according to the immune infiltrating cells in these two clusters, as showed in **Figure 2d**, revealed a positive correlation between neutrophils, eosinophils, B-cells, macrophages, memory B cells, mast cells and recurrence, a negative correlation between CD4+ T cells (including CD4+ effector memory T cells, Tems, and CD4+ central memory T cells, Tcms), CD8+ T cells (including CD8+ Tems and CD8+ Tcms), regulatory cells (Tregs), NK cells, monocyte, plasmacytoid dendritic cells (pDCs) and recurrence. These data suggested that CD4+ T cells (including CD4+ Tems and CD4+ Tcms), CD8+ T cells (including CD8+ Tems and CD8+ Tcms), Tregs, NK cells, monocyte, pDCs may had a beneficial effect on the prognosis of GISTs.

**Figure 2.**
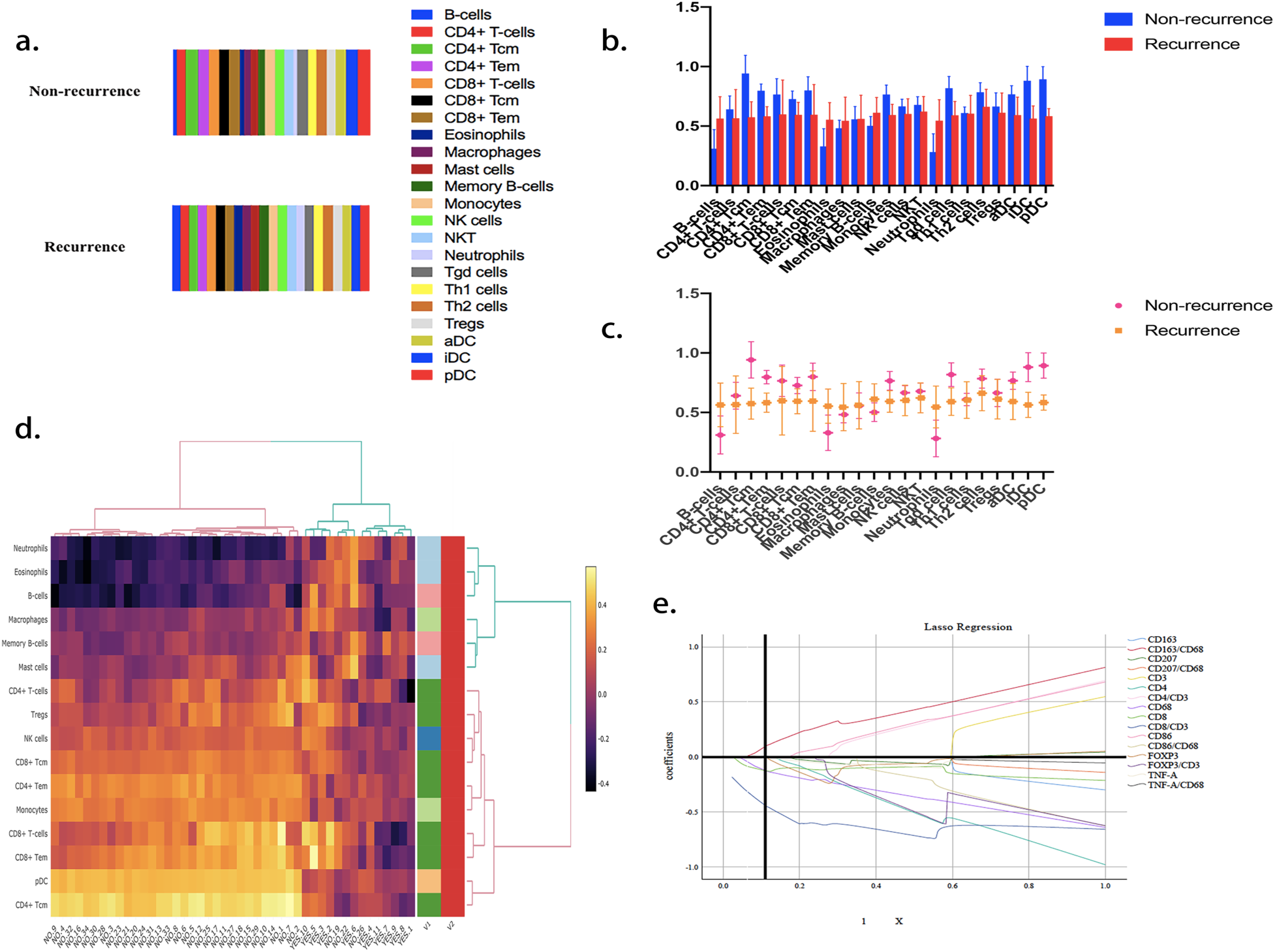
a. Relative fractions of 22 immune cell types in the samples from the training cohort in clusters with non-recurrence and clusters with recurrence; b and c. Comparison of relative fractions of the immune cell types in the two clusters; d. Correlation matrix followed by unsupervised hierarchical clustering in immune cell fractions of GEO dataset; e. LASSO coefficients profiles of included immune features.

The LASSO Cox regression model was performed to select the most useful prognostic immune features on the basis of RFS (**Figure 2e**). Finally, a set of 4 immune features based on their expression data along with their coefficients weighted by the LASSO Cox model were identified and included into a risk score formula. Risk score = (30.64 × the level of CD8) + (35.496 × the level of CD68) + (4.848 × the level of (CD8/CD3)) – (0.198 × the level of (CD163/CD68)).

### Patient characteristics of the validation cohort

To validate the performance of the risk score, 54 patients with locally primary GISTs who underwent complete surgery were included in the validation set. Detailed clinical characteristics of included patients in the validation set are summarized in **Table 1**. The validation set included 34 males and 20 females, with median age at diagnosis being 60 years (range from 35 to 83 years). The majority of tumors were located in the stomach, only 27.8% of patients (15/54) were located in the small intestine, 3.7% (2/54) in the colon. After a follow-up of 60 months, 29.6 percent of patients (16/54) developed recurrence. The 3- and 5-year RFS were 87.0% and 70.4%, respectively.

**Table 1.** Detailed clinical characteristics of the patients in the validation cohort

| Variables | Total Number | Recurrence(%) | Non-recurrence(%) |
| --- | --- | --- | --- |
| Sex |  |  |  |
| male | 34 | 9(56.3%) | 25(65.8%) |
| female | 20 | 7(43.7%) | 13(34.2%) |
| Age (years) |  |  |  |
| ≤60 | 28 | 10(62.5%) | 18(47.4%) |
| >60 | 26 | 6(37.5%) | 20(52.6%) |
| Tumor size (cm) |  |  |  |
| ≤2 | 2 | 0(0.0%) | 2(5.3%) |
| >2, ≤5 | 16 | 0(0.0%) | 16(42.1%) |
| >5, ≤10 | 20 | 4(25.0%) | 16(42.1%) |
| >10 | 16 | 12(75.0%) | 4(10.5%) |
| Mitotic count (/50HPF) |  |  |  |
| ≤5 | 24 | 3(18.8%) | 21(55.3%) |
| 6-10 | 13 | 4(25.0%) | 9(23.7%) |
| >10 | 17 | 9(56.2%) | 8(21.0%) |
| Tumor location |  |  |  |
| gastric | 37 | 5(31.3%) | 32(84.2%) |
| small intestine | 15 | 11(68.7%) | 4(10.5%) |
| colon | 2 | 0(0.0%) | 2(5.3%) |
| Tumor rupture |  |  |  |
| no | 50 | 13(81.3%) | 37(97.4%) |
| yes | 4 | 3(18.7%) | 1(2.6%) |
| Tumor necrosis |  |  |  |
| no | 47 | 12(75.0%) | 35(92.1%) |
| yes | 7 | 4(25.0%) | 3(7.9%) |
| Histologic type |  |  |  |
| spindle | 51 | 16(100.0%) | 35(92.1%) |
| epithelioid | 2 | 0(0.0%) | 2(5.3%) |
| mixed | 1 | 0(0.0%) | 1(2.6%) |
| CD117 |  |  |  |
| positive | 54 | 16(100.0%) | 38(100.0%) |
| negative | 0 | 0(0.0%) | 0(0.0%) |
| DOG-1 |  |  |  |
| positive | 52 | 16(100.0%) | 36(94.7%) |
| negative | 2 | 0(0.0%) | 2(5.3%) |
| CD34 |  |  |  |
| positive | 44 | 14(87.5%) | 30(84.2%) |
| negative | 10 | 2(12.5%) | 8(21.0%) |
| Ki-67 |  |  |  |
| <10% | 39 | 10(62.5%) | 29(76.3%) |
| ≥10% | 15 | 6(37.5%) | 9(23.7%) |
| Genotype |  |  |  |
| exon 9 | 6 | 4(25.0%) | 2(5.3%) |
| exon 11 | 15 | 3(18.7%) | 12(31.6%) |
| exon 12 | 4 | 1(6.3%) | 3(7.9%) |
| exon 18 | 4 | 0(0.0%) | 4(10.5%) |
| NA | 25 | 8(50.0%) | 17(44.7%) |

### Immune features based on CD8, CD68, CD8/CD3, CD163/CD68 predicted recurrence of GISTs

According to previous bioinformatics analyses, it could be concluded that these four immune features (CD8, CD68, CD8/CD3, CD163/CD68) may had a significant influence on prognosis of GISTs. We then detected the expression of CD8, CD3, CD68, CD163 in the infiltration of tumor. Typical IHC staining of CD8, CD3, CD68, CD163 were showed in **Figure 3**, respectively. The level of immune features was identified according to the number of positively stained cells in intra-tumoral tissues of each patient. The mean value of 5 representative fields for each marker were scored, recorded for further analyses. We take the value higher than the median as high, lower than the median as low. In the validation cohort, clusters with non-recurrence had an obviously higher expression of CD8, CD68, while a lower expression of CD163 compared with clusters with recurrence (**Figure 4a**). According to the survival analysis showed in **Figure 4b**, patients with high CD8, CD68, CD8/CD3 expression had higher 5-year RFS compared to those with low CD8, CD68, CD8/CD3 expression. While patients with high CD163/CD68 expression were inversely related to 5-year RFS compared to those with low CD163/CD68 expression. These results were proved to be consistent to the risk score mentioned above.

**Figure 3.**
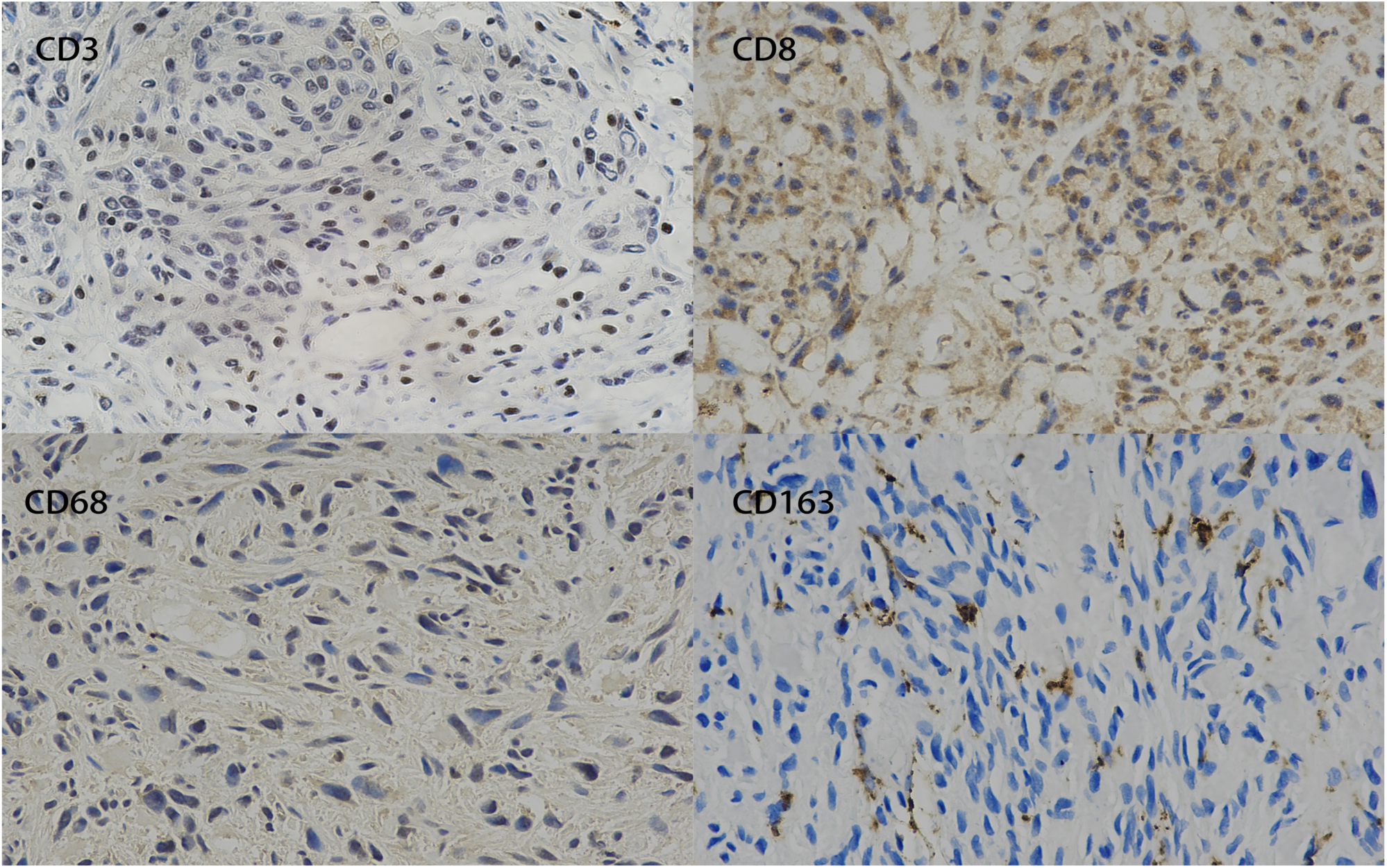
Typical immunohistochemical staining of CD8, CD3, CD68, CD163.

**Figure 4.**
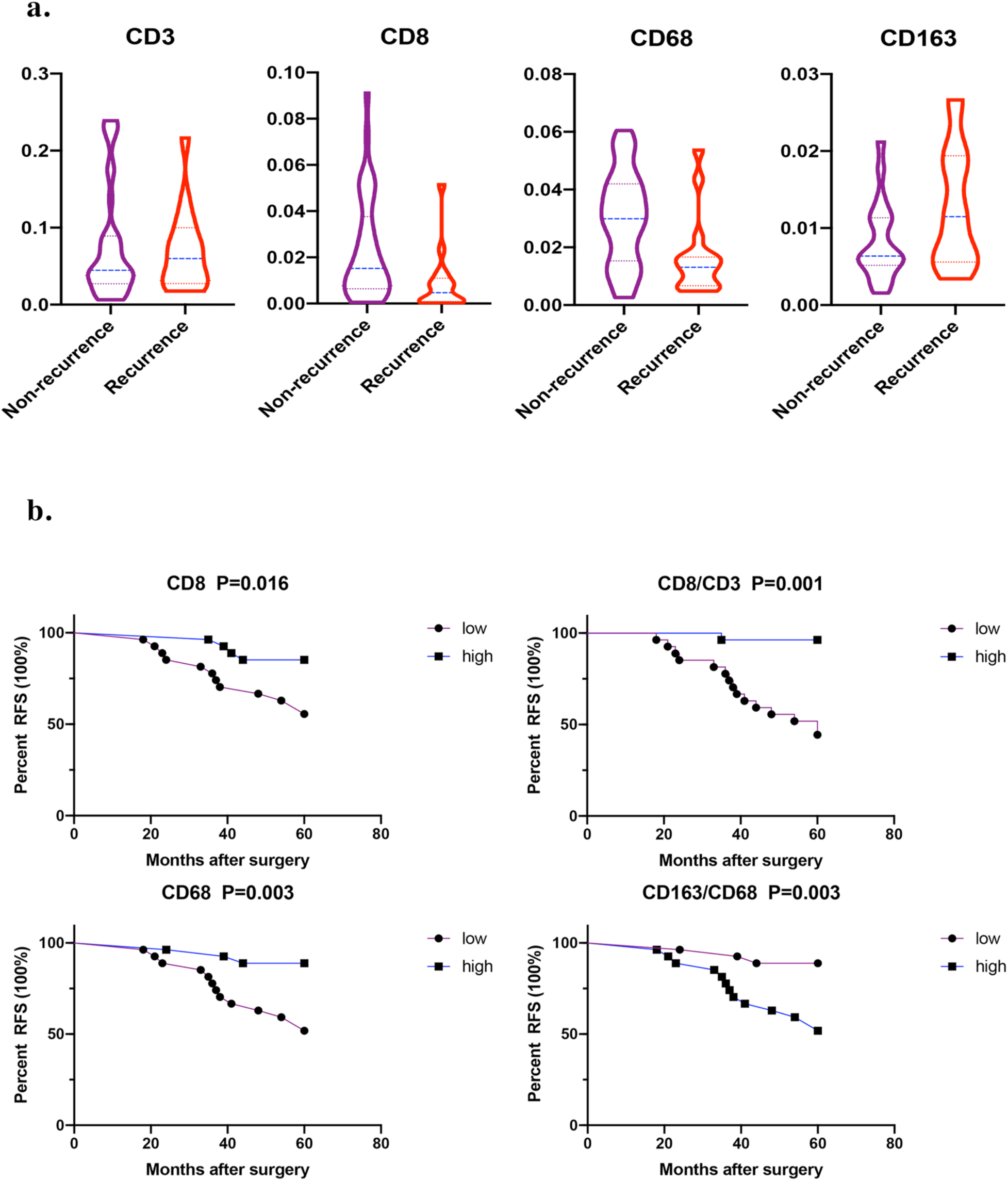
a. Comparison of the level of CD8 (*p*=0.02), CD3 (*p*=0.76), CD68 (*p*=0.01), CD163 (*p*=0.01) expression in tumors with recurrence and tumors with non-recurrence; b. Kaplan-Meier survival curve of the validation cohort categorized by different levels of CD3, CD8/CD3, CD68, CD163/CD68 expressions (high versus low).

### The IPC stratified patients into groups of different risk

We calculated the exact value of the risk score according to the formula mentioned above. For the convenience of further statistics, we take the reciprocal of the risk score as the final prognostic classifier (IPC). IPC =1/{(30.64 × the level of CD8) + (35.496× the level of CD68) + (4.848 × the level of (CD8/CD3)) – (0.198 × the level of (CD163/CD68))}. We take the IPC lower than the median as low-immune risk group, higher than the median as high-immune risk group in further analyses. As showed in **Figure 5a**, patients were significantly stratified into two groups of which the low-IPC group were associated with longer 5-year RFS compared high-IPC group (92.6% vs 48.1%, *p* < 0.001), revealing IPC a potential useful prognostic classifier for GISTs. Further univariate and multivariate analysis also demonstrated that IPC, as well as tumor size, mitotic count and rupture, were independent predictors of RFS for the validation cohort (**Table 2**). In addition, in subgroup of high-risk according to the modified NIH risk classification, the IPC still split patients into two groups (5-year RFS,low-IPC group vs high-IPC group, 85.7% vs 30.0%, *p* =0.002), which was showed in **Figure 5b**. The ability of the IPC to predict the recurrence of GIST was shown to have an area under the curve (AUC) of 0.842 in the validation cohort (**Figure 5c**). Meanwhile, the DCA curve for the IPC presented in **Figure 5d** showed the IPC a higher net benefit than both “treat all” or “treat none” methods for the threshold probability within a range of 0-0.62.

**Table 2.** Univariate and multivariate analysis for RFS of patients with GISTs

|  | Univariate analysis |  |  | Multivariate analysis |  |  |
| --- | --- | --- | --- | --- | --- | --- |
|  | HR | 95%CI | *p* value | HR | 95%CI | *p* value |
| Gender | 1.350 | 0.503-3.628 | 0.554 |  |  |  |
| Age | 1.005 | 0.961-1.050 | 0.840 |  |  |  |
| Tumor size | 1.240 | 1.136-1.353 | <0.001 | 1.217 | 1.064-1.392 | 0.004 |
| Tumor site | 6.928 | 2.398-20.012 | <0.001 | 0.850 | 0.159-4.552 | 0.849 |
| Mitotic count | 1.055 | 1.019-1.093 | 0.003 | 1.108 | 1.054-1.165 | <0.001 |
| Tumor rupture | 6.918 | 1.923-24.882 | 0.003 | 11.535 | 1.937-68.674 | 0.007 |
| Tumor necrosis | 2.338 | 0.754-7.251 | 0.141 |  |  |  |
| Ki-67 | 1.044 | 0.971-1.123 | 0.245 |  |  |  |
| Histologic type | 0.097 | 0.000-79.747 | 0.496 |  |  |  |
| IPC | 2.739 | 1.742-4.307 | <0.001 | 3.521 | 1.553-7.981 | 0.003 |

**Figure 5.**
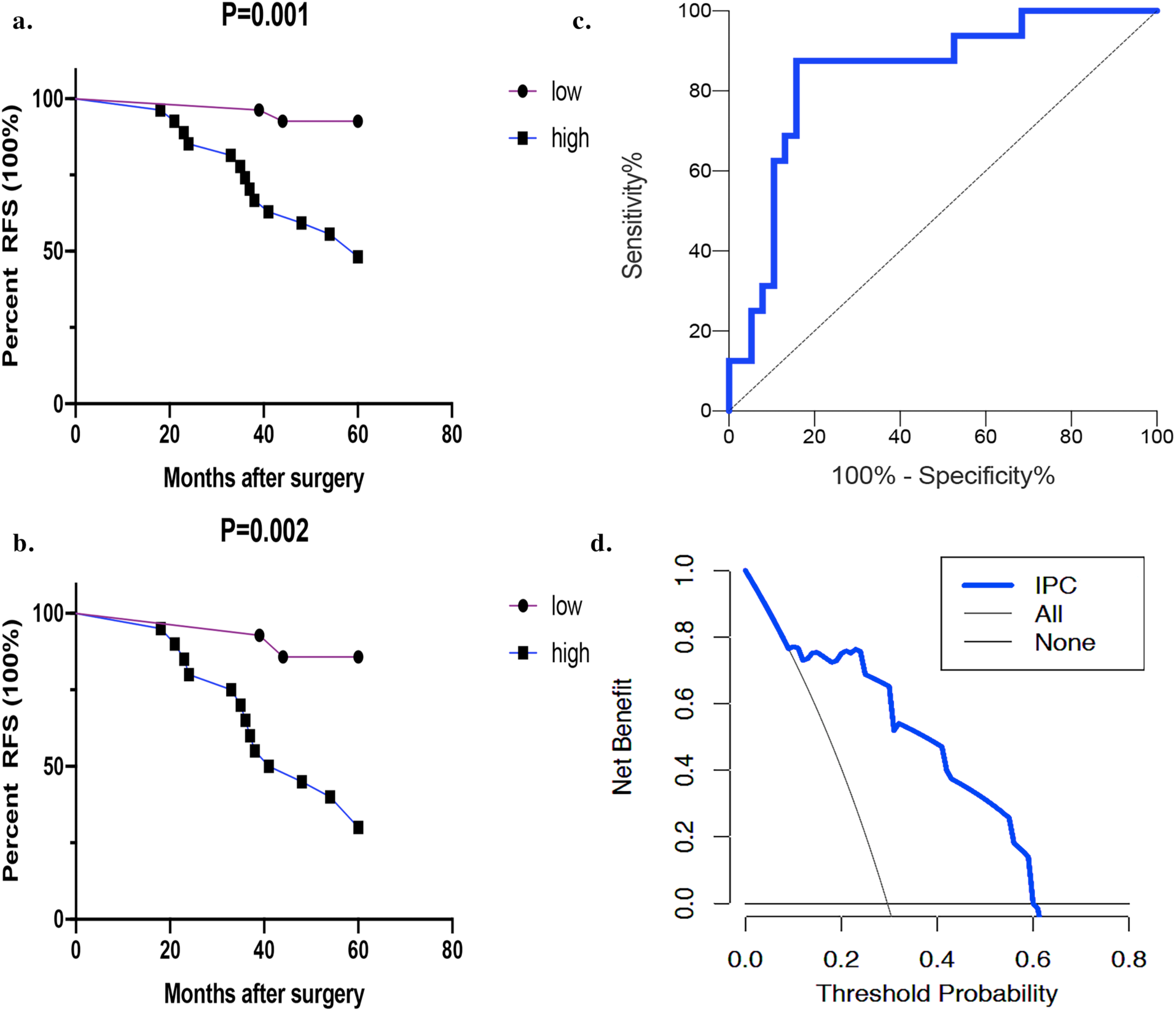
a. Kaplan-Meier survival curve of the whole validation cohort categorized by IPC; b. Kaplan-Meier survival curve of the validation cohort categorized by IPC of high risk group of the modified National Institutes of Health (NIH) classification; c. ROC analysis of the ability of the IPC to predict the recurrence of GISTs; d. Decision curve analysis evaluating the clinical utility of the IPCGIST showed that the IPC had a higher net benefit than both “treat all” or “treat none” methods for the threshold probability within a range of 0-0.62.

### Comparison of the predicting accuracy of IPC with other criteria

To verify the predicting accuracy of the IPC compared with the modified NIH risk classification and prognostic heat maps by *Joensuu*, ROC analysis was performed as showed in **Figure 6**. The IPC (AUC 0.842) provided a more accurate prognosis than the modified NIH criterion (AUC 0.763), while less accurate than heat maps by *Joensuu* (AUC 0.929).

**Figure 6.**
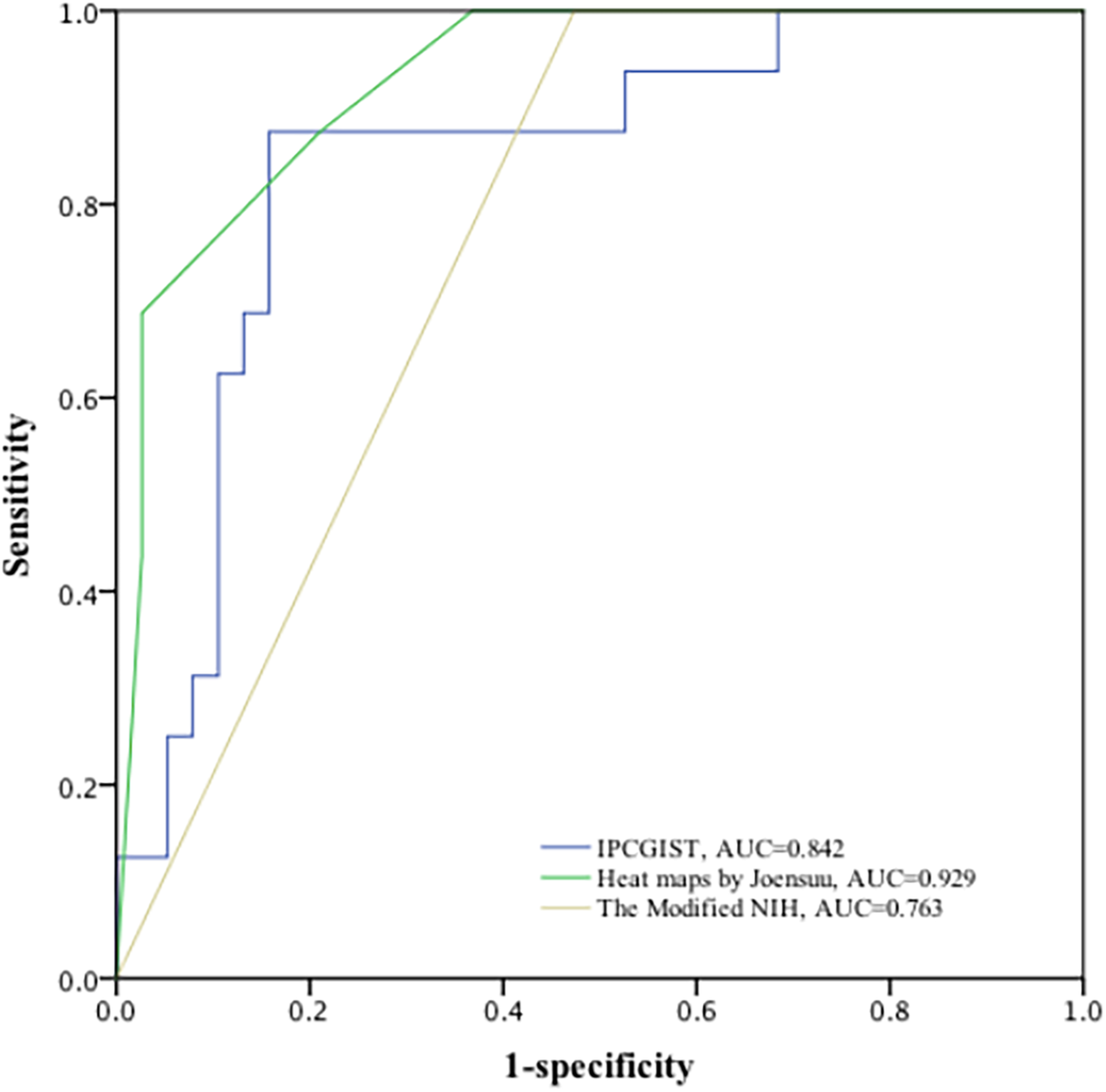
Comparison of the predicting accuracy of the IPC, the modified NIH classification and heat maps by *Joensuu* in the validation set.

## Discussion

Significant prognostic heterogeneity has been described in GISTs with recurrence risk varying from almost none (for the smallest GISTs) to well over 50% after complete resection.^22^ Accurate assessment of prognostic is especially important for the selection of adjuvant imatinib therapy.^23^ Several validated classification criteria to predict risk of recurrence after complete surgery have been proposed and been proved to provide useful but incomplete information.^18,19,24–27^ In routine clinical risk prognostic risk classifications, tumor size, tumor site, mitotic count and rupture were key prognostic determinants in GISTs for clinical workers and patients.^18,19,26^ However, prognostic information these current clinical prognostic risk classifications afforded was found to be limited. Accumulating evidence supposed that the progression of GIST is affected not only by its intrinsic characteristics, but also by extrinsic immune effectors.^6^ Therefore, we established a classifier named IPC based on 4 immune features as a novel prognostic tool which was independent of these classical predictors as above mentioned to help to better stratify patients and afford additional information when considering adjuvant imatnib therapy. Though the IPC was constructed based on genomic data from GEO dataset, subsequent IHC of the including four immune features in the validation set confirmed its prognostic value. It was straightforward through identification of tumor infiltrating immune features with IHC and could be a procedure clinically applicable. Besides, the recurrence predictive ability of our IPC seemed a little higher than the modified NIH classification according to ROC analysis, but lower than the heat maps by *Joensuu*. Although the modified NIH classification is crucial to establish a treatment strategy and the heat maps by *Joensuu* afford better accurate prognosis of GISTs, both of them were performed only on the basis of its anatomical information. Instead, our IPC could indicate the internal immunological characteristics of GISTs and afford completely different information from the routine clinical prognostic criteria. In this study, the integration of various immune features seemed reinforced the prognostic ability of the modified NIH classification, thereby adding prognostic value to the modified NIH classification. Actually, in the validation set, patients of the high-risk group according to modified NIH classification were further split in two subgroups, of which subgroup with relative low-IPC was related to a statistically significant longer RFS, which might afford additional prognostic information when considering adjuvant imatinib therapy.

Early IHC in human GIST has proved the presence of infiltrating CD3+ T cells, CD8+ T cells and macrophages.^7,28^ In the current study, CD8, CD8/CD3, CD68, CD163/CD68 were the main four immune features included in the novel immune-based classifier. IHC of these markers in the validation set showed that GISTs with high CD8 expression had better RFS compared with those with low CD8 expression. Similarly, previous trial by *Balachadran* and collegues have found that mice lacking T and B cells had larger tumors than control group, while mice lacking B cells did not.^29^ Further they found that imatinib-naïve GIST mice depleted for 4 weeks of CD8+ T cells had larger tumors, indicating CD8+ T cells favorable prognostic predictor in GISTs. The quantity of CD3+ T cells has been found be correlated with smaller tumor size, reduced recurrence and increased survival of GISTs.^6^ However, our present study showed a positive correlation between the tumor behavior and CD8/CD3, not CD3, possibly indicating the more important role of CD8+ T cells in TILs of GISTs. It has been reported for decades that infiltration of CD8+ T cells could promote the prognosis of other human cancers^30,31^ and be proved as an independent favorable predictor for survival in several cancers, ovarian cancer,^32,33^ renal cell carcinoma,^34^ nonsmall cell lung cancer, ^35^ breast cancer,^36,37^ for example. Especially in colorectal cancer, the prognostic value of CD8+ lymphocytes (combined with CD3+ and CD45RO+ lymphocytes) was found to be even superior to the Union for International Cancer Control (UICC) -TNM staging system.^31^ Cytotoxic CD8+ T cells, which act as crucial components of tumor-specific cellular adaptive immunity, could attack tumor cells that presented tumor-related antigen peptide with major histocompatibility complex class I on surface ^38,39^ and produce interferon gamma, inducing tumor cell killing through cell inhibition, apoptosis, and induction of macrophage tumoricidal activity.^40^ Besides, our study also showed a positive relation between CD68 expression and RFS, while a negative relation for CD163/CD68, of which CD163 was predominantly expressed in M2 phenotype macrophages known to play a role in immune suppression, indicating a potential relationship between macrophages and the prognosis of GISTs. Macrophages were used to be recognized as evolutionarily ancient cells involved in tissue homeostasis and immune defense against pathogens, now rediscovered as regulators of several diseases.^41^ Depletion of TAMs was found to be related with retirement of cytotoxic T cell reactions and suppression of tumor growth.^41^ However, the correlation between the number of TAMs and the clinical behavior of GISTs is not clear.^42^ Even so, interestingly, TAMs were found to be the numerous infiltrating immune cells in GISTs, and metastatic GISTs harbored twice as much M2 type macrophages as primary lesions.^7^ Previously, *Maud Toulmonde* and colleagues observed prominently infiltration of CD163+ macrophages in GISTs, contributing to immune-suppression and primary resistance to PD-1 inhibition,^11^ which may help to elucidate the observed negative correlation between CD163/CD68 expression and prognosis of GISTs in this research.

Mutations in type III receptor kinases account for 85% of patients, of which the majority respond to treatment with the tyrosine kinase inhibitor (TKI).^43–45^ While, part of GISTs were found to harbor mutations resistant to currently TKIs, which highlighted a increasing urgency in developing novel methods to overcome the resistance of TKIs. The introduction of immunotherapy with the monoclonal antibodies against the immune checkpoints PD-1 and CTLA-4 played a revolutionized role in the development of the cancer treatment.^46,47^ With evolving understanding of interaction between the immune environment and the tumor in GISTs, combinative therapy with imatinib and PD-1 blockade was found to show effects superior to treatment with imatinib alone in previous clinical models,^29,48^ while TKI combined with CTLA-4 blockade reported no synergistic activity in GISTs.^49^ Our study provides an opportunity and rationale for developing novel effective forms of immunotherapy. In our study, a positive relation between survival and CD68 expression and the negative relation between survival and CD163/CD68 suggest that depletion of TAMs may be detrimental, in consistent with previous research,^50^ while changing the polarization status of TAMs might be a promising supplementary approach for treatment of GIST in the future. Indeed, the tumor weight increased after four weeks of TAMs depletion with the small molecule CSF1R inhibitor PLX5622 in GIST mice.^51^ However, after 2 weeks of anti- CD40 treatment, the tumor infiltrating TAMs were more M1-like, and the combined treatment with imatinib and anti-CD40 achieved greater direct tumor inhibition in vitro in GIST.^52^ Besides, inhibiting STAT3-induced gene expression through blocking ERK5 was found to be a potential effective treatment for cancer via reprogramming macrophages towards an antitumor state.^53^ There remained some limitations in this study to be discussed. First, it was a retrospective research with its inherent defects, and with limited generalizability because of the relative small samples from both the GEO dataset and our validation cohort as the rarity of this disease. The second limitation is that the immune features included in this study were incomplete. However, according to previous studies, the common infiltrating immune cells were absolutely included and we think the difference caused by this deficiency could be ignored. Third, we failed to further evaluate the model stability in different clinical groups because of the limited candidates in this study. Thus, a prospective, international, multicenter clinical trial with large sample is in need to further validate our findings. Besides, the biological mechanisms about how the immune markers are involved in GISTs remain to be investigated in further researches. In addition, studies about whether the IPC could predict the responses of GISTs to immunotherapy should be conducted in the future. Finally, the IPC was constructed and validated based on patients without imatinib adjuvant therapy, which may not be suitable for patients with imatinib therapy.

In conclusion, the IPC could probably be an effective tool to predict RFS after complete resection of GISTs, and afford additional prognostic value to the routine clinical prognostic criteria for GISTs. Thus, the IPC might be applicated in clinical process such as patient counseling, decision regarding personalized adjuvant imatinib therapy, and follow-up scheduling in the future. In addition, our study affords rational of possibility of immunotherapy in GISTs in a sense. Prospective studies are in an urgent need to further validate the analytical accuracy of the IPC in assessing prognosis for individual management of GISTs.

## Data Availability

The data referred to in the manuscript was available from the GEO dataset.

## Abbreviations

GIST: gastrointestinal stromal tumor
IPC: immune-based prognostic classifier
RFS: recurrence free survival
TAMs: tumor-associated macrophages
Tems: effector memory T cells
Tcms: central memory T cells
Tgd cells: T Gamma/Delta cells
TILs: tumor infiltrating lymphocytes
Tregs: regulatory cells
NIH: National Institutes of Health
UICC: the Union for International Cancer Control
AUC: area under the curve
PD-L1: programmed death-ligand1
NK: natural killer
LASSO: least absolute shrinkage and selection operator method
HR: hazard ratio
CI: confidence interval
ROC: receiver operating characteristic
DCA: decision curve analysis

## Acknowledgments

The authors thank Yuan Tang, Huijiao Chen, Hongying Zhang, Zhang Zhang, and Ni Chen from Pathology Department of West China Hospital, Sichuan University for supporting this work.

## Declaration of interest statement

The authors declare no conflicts of interests.

## Table legends

Table 1. Detailed clinical characteristics of the patients in the validation set

Table 2. Univariate and multivariate Cox regression analysis for RFS of patients with GISTs

## Notes

### Competing Interest Statement

The authors have declared no competing interest.

### Funding Statement

This work was supported by Chinese Medical Board Grant on Evidence-Based Medicine, New York, USA (No. 98-680), National Natural Science Foundation of China (No. 30901427)

